# Bloody jello: Colloidal physics states acanthocyte sedimentation rate as a diagnostic biomarker for neuroacanthocytosis

**DOI:** 10.1101/2020.09.01.20185041

**Authors:** Alexis Darras, Kevin Peikert, Antonia Rabe, François Yaya, Greta Simionato, Thomas John, Anil Kumar Dasanna, Semen Buvalyy, Jürgen Geisel, Andreas Hermann, Dmitry A. Fedosov, Adrian Danek, Christian Wagner, Lars Kaestner

## Abstract

Chorea-acanthocytosis and McLeod syndrome are the core diseases among the group of rare neurodegenerative disorders called neuroacanthocytosis syndrome (NAS). NAS patients have irregularly spiky erythrocytes, so-called acanthocytes. Their detection is a crucial but error-prone parameter in the diagnosis of NAS, often leading to misdiagnosis. Based on the standard Westergren method, we show that the acanthocyte sedimentation rate (ASR) with a two-hour read-out is significantly prolonged in Chorea-acanthocytosis and McLeod syndrome without overlap compared to the erythrocyte sedimentation rate (ESR) of controls. Thus, the ASR/ESR is a clear, robust and easily obtained diagnostic marker. Mechanistically, through modern colloidal physics, we show that acanthocyte aggregation and plasma fibrinogen levels slow down the sedimentation. This study is also a hallmark of the physical view of the erythrocyte sedimentation by describing anticoagulated blood in stasis as a percolating gel, allowing the application of colloidal physics theory.

## 1 Introduction

Neuroacanthocytosis syndromes (NASs) are a group of rare genetic neurodegenerative diseases, the two core disorders being chorea-acanthocytosis (ChAc, OMIM #200150) and McLeod syndrome (MLS, OMIM #300842)^1–7^. While both diseases are characterized by neurological symptoms such as hyper- and hypokinesia, epileptic seizures and cognitive impairment, ChAc usually manifests much earlier in life (in the twenties) than MLS (usually after 40 years of age). The mean life expectancy after diagnosis has been reported to be approximately 11 years for ChAc and 21 years for MLS, leading to premature death in most cases^8^. The courses of the diseases are heterogeneous, usually slowly progressive and often result in severe disability, while treatment options remain purely symptomatic so far^9^. Diagnosis is confirmed by genetic testing (VPS13A gene in ChAc, XK gene in MLS) and chorein in Western blot (ChAc) or immunohematological assessment (MLS)^5^, although the initial detection often relies on blood smears^10^. One of the common features of NAS is the presence of acanthocytes among erythrocytes^4–6^. Acanthocytes are deformed erythrocytes whose irregular membrane presents disordered, asymmetric spikes.

For many of these rare diseases, diagnosis is challenging, and misdiagnosis is frequent, which additionally increases the patients’ burden of disease. One of the reasons for delayed diagnosis is related to the difficult detection of acanthocytes in classical blood smears and routine laboratory testing: sensitive detection requires special isotonically diluted blood samples^10^, the number of acanthocytes varies among the patients, and their similarity to echinocytes often makes them hard to identify^6^. Therefore, it would be desirable to have an easier and more reliable initial diagnostic biomarker.

Here, we proposed the erythrocyte sedimentation rate (ESR) as a convenient, inexpensive and objective parameter that allows routine and systematic tests to be used to diagnose NAS. At the same time, we investigated the physical cause of the change in the acanthocyte sedimentation rate (ASR). In doing so, we devised a novel physical view of the sedimentation process based on colloidal physics that is in good agreement with the experimental data.

## 2 Results

### Significance of the ESR in Neuroacanthocytosis Syndrome Patients

Time-resolved measurements of the ESR following the standard Westergren method were performed on 6 ChAc patients, 3 MLS patients and 8 healthy controls. The major hematological data as well as the neurological phenotypes of the individuals are given in Table 1. An image of the sedimentation tubes, the time curves of representative ASR/ESR measurements, the statistical presentation of patient-based data at selected time points and the overview of all measurements at selected time points are summarized in Fig. 1A-D, respectively.

**Table 1:** Overview of the patients and controls

| subjects | sex | age range (y) | hematocrit (%) | hemoglobin (g/dl) | RBC number ($10^{12}/l$) | main clinical characteristics |
| --- | --- | --- | --- | --- | --- | --- |
| ChAc-1 | male | 50-55 | 43 | 15.1 | 4.67 | Parkinsonim, dystonia, dysarthria, peripheral neuropathy,depression |
| ChAc-2 | female | 50-55 | 40 | 13.8 | 4.21 | Epilepsy,parkinsonism dystonia, dysarthria, peripheral neuropathy, cognitive impairment |
| ChAc-3 | male | 50-55 | 40 | 14.2 | 4.15 | Epilepsy, Parkinsonism, dystonia, dysarthria, dysphagia, peripheral neuropathy, cognitive impairment |
| ChAc-4 | male | 30-35 | 46 | 16.2 | 5.40 | Drug resistant epilepsy, mild chorea, tics, cognitive impairment, peripheral neuropathy, myopathy |
| ChAc-5 | male | 25-30 | 43 | 15.7 | 4.91 | Drug resistant epilepsy, mild chorea, tics, cognitive impairment, irritability, anxiety, depression, psychosis |
| ChAc-6 | male | 40-45 | 41 | 15.2 | 4.96 | Epilepsy, feeding dystonia, orofacial dyskinesia, chorea, peripheral neuropathy, myopathy, impulse control disorder |
| MLS-1 | male | 55-60 | 43 | 16.0 | 4.82 | Myopathy |
| MLS-2 | male | 50-55 | 42 | 15.3 | 4.67 | Epilepsy, peripheral neuropathy, myopathy |
| MLS-3 | male | 50-55 | 45 | 16.2 | 5.17 | Neuropathy,myopathy |
| female controls (average, N=4) |  | 64±10 | 38.5±2.6 | 12.6±1.3 | 4.36±0.19 | — |
| male controls (average, N=4) |  | 38±11 | 43.4±2.1 | 15.5±0.8 | 5.17±0.1 | — |

**Figure 1.**
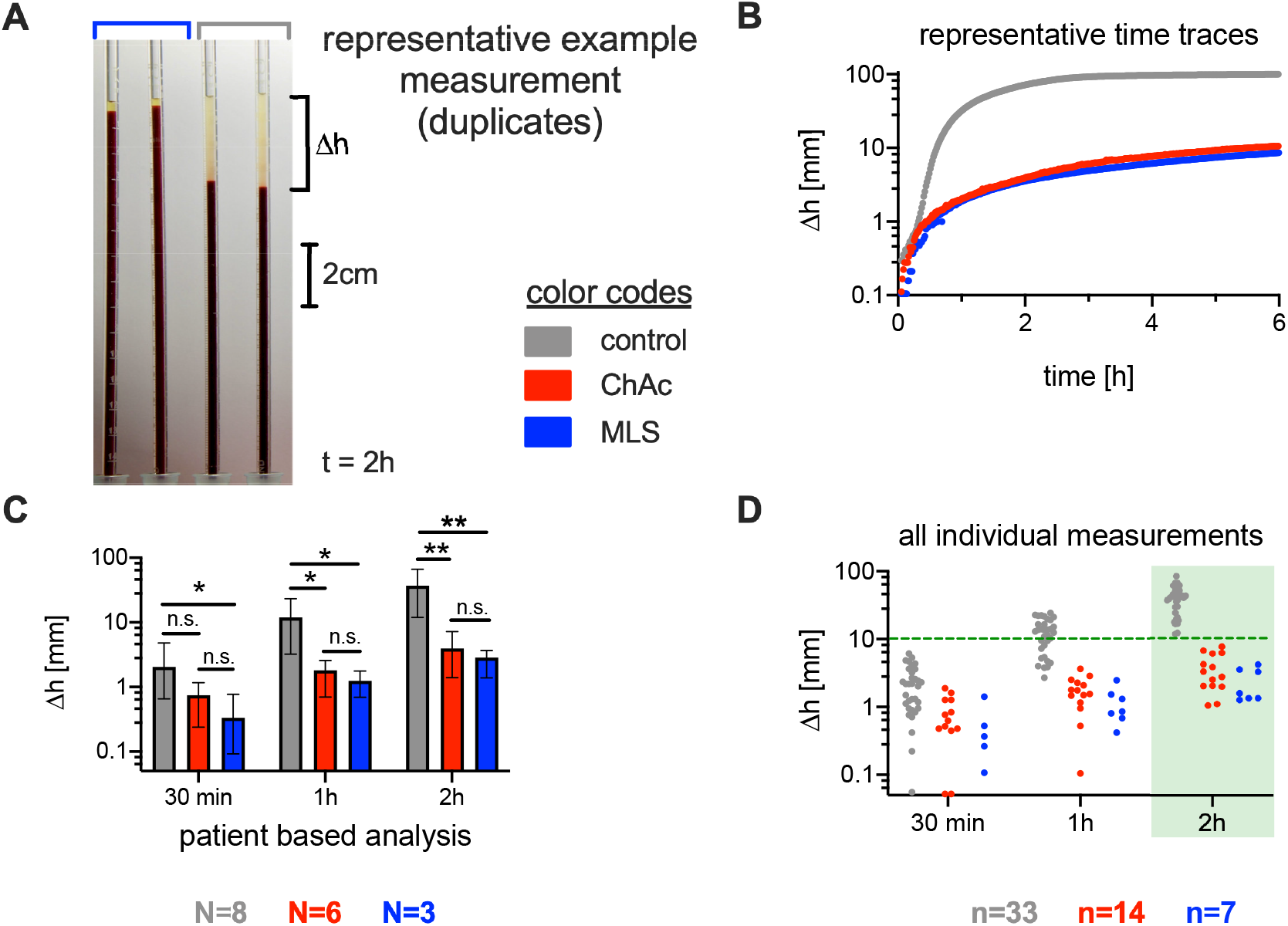
Comparison of the ESR between neuroacanthocytosis patients and healthy controls. A: ESR measurement setup: standard Westergren tubes are filled with full blood and left to rest. The sedimentation height is measured over time. The picture was taken after 2 h. The first two tubes contain blood from an MLS patient (MLS-3), and the last two tubes contain blood from a healthy control donor. B: Representative time traces of the sedimentation height for the different conditions. C: Statistics on sedimentation height after typical times for healthy controls, ChAc patients and MLS patients. The bar height represents the average value, and the error bar depicts the range of individual values. Stars indicate the significance levels (Brown-Forsythe and Welch ANOVA test): n.s., not significant (p-value*>*0.05); * p*<*0.05; and ** p*<*0.01. N refers to the number of patients. D: Span of all individual sedimentation height measurements (each blood sample was measured at least in duplicate, and the blood of some patients was measured at different time points). After 2 h, a Δ*h* of 10 mm could be regarded as a threshold to differentiate the ChAc and MLS conditions. n refers to the number of individual measurements.

Although the number of patients was limited, at 30 min, the difference in ESR was significantly lower for MLS patients than for controls (*p* = 0.028). At 1 h, we found a significant difference between both ChAc patients (*p* = 0.022) and MLS patients (*p* = 0.017) and healthy controls. After 2 h, the p-value decreased to 0.006 for ChAc patients and to 0.005 for MLS patients relative to the healthy controls (Fig. 1C).

However, a clear systematic difference between the whole range of measurements is only reached after 2 h. A sedimentation height of 10 mm can be regarded as a threshold (green dashed line in Fig. 1D), i.e. only if the sedimentation phase is less than 10 mm within 2h, ESR indicates ChAc and MLS. These results are sex independent (cp. Supplemental Fig. 1A).

### Comparison of the Acanthocyte Percentage with the ESR

To date the diagnosis of NAS has involved the detection of acanthocytes. Consequently, here blood smears were prepared based on the protocol from Storch *et al*.^10^ and later stained according to the Pappenheim method (see Supplemental Data). An example is presented in Fig. 2A. The manual count of cell shapes is prone to be subjective, biased and inaccurate and therefore reproducible with only limited reliability, as outlined in Supplemental Fig. 2. For a better assessment, we fixed fresh cells in 0.1 % glutaraldehyde and performed confocal imaging of *z*-stacks to 3D render the erythrocytes. See Supplemental Data for detailed protocol. Figure 2B shows a number of representative acanthocytes in 3D, which were used for further analysis.

**Figure 2.**
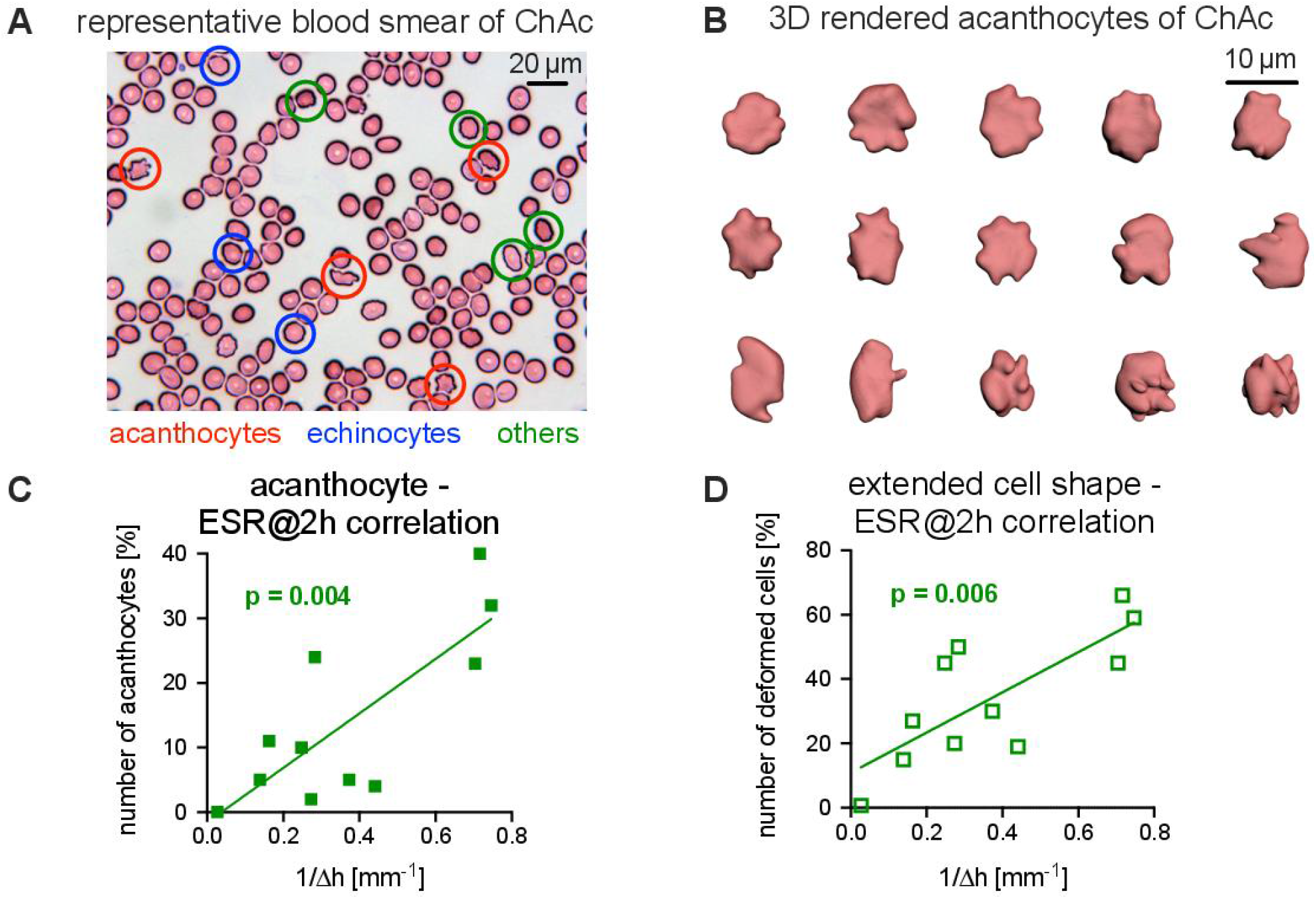
Relation of acanthocyte counts to ESR. A: A representative stained blood smear of a ChAc patient. B: Representative 3D-rendered acanthocytes from the freshly fixed blood of a ChAc patient. C: Plot of the number of acanthocytes vs. the inverse of the ESR at 2 h and the corresponding correlation analysis. D: Plot of the number of all deformed erythrocytes (echinocytes, acanthocytes and other unspecified cell shapes) vs. the inverse of the ESR at 2 h and the corresponding correlation analysis. Panels C and D contain measurements from 9 patients (cp. Table 1), where 1 patient was measured twice (independent blood sampling on different days). Since none of the controls presented with acanthocytes, they are summarized in one data point with their average ESR. The plotted line represents the linear regression. The analysis reveals a significant correlation for both parameters (panels C and D, *R*^2^ coefficient of 0.61 and 0.59, respectively).

We observed that the ESR tends to decrease when the fraction of acanthocytes increases. To relate the ESR to the number of acanthocytes, the inverse of the ESR at 2 h is plotted as a function of the number of acanthocytes (Fig. 2C). In a more general approach, we plotted the inverse of the ESR at 2 h against the number of all deformed cells (echinocytes, acanthocytes and others; Fig. 2D). The analysis revealed a significant correlation for both parameters (*R*^2^ coefficient of 0.61 and 0.59, respectively). Consistent with this finding, the p-value decreased from 0.037 (acanthocytes) to 0.011 (all deformed cells).

### Explanations for Differences in the ESR

Because there was no significant difference in the ASR between ChAc and MLS patients (Fig. 1), we do not discriminate between ChAc and MLS patients for further statistical comparisons. Therefore, we subsume them into one NAS group.

The reasons for the differences in the ESR are varied and can depend on the erythrocytes, the blood plasma or the erythrocytes-plasma volume relation (i.e. the hematocrit or erythrocyte volume fraction *ϕ*). The last reason can be excluded since there was no significant difference in the hematocrit between the groups (cp. Table 1 and Supplemental Fig. 1B). The primary physical parameters such as the density of the individual erythrocytes, represented by the mean cellular hemoglobin concentration (MCHC), as well as the constitution of the plasma can also be part of the explanation. However, the MCHC indicates a higher density of erythrocytes in the NAS patients (Supplemental Fig. 3C), a feature that would favor faster sedimentation and therefore rules it out as an explanation. In contrast, the total plasma protein analysis (Supplemental Fig. 4A), as a representative of the plasma content, indicates a lower protein concentration and therefore a first hint for a plasma contribution to the ESR change. In the following sections, we present a more detailed view of the influence of both blood components, erythrocytes and plasma.

**Figure 3.**
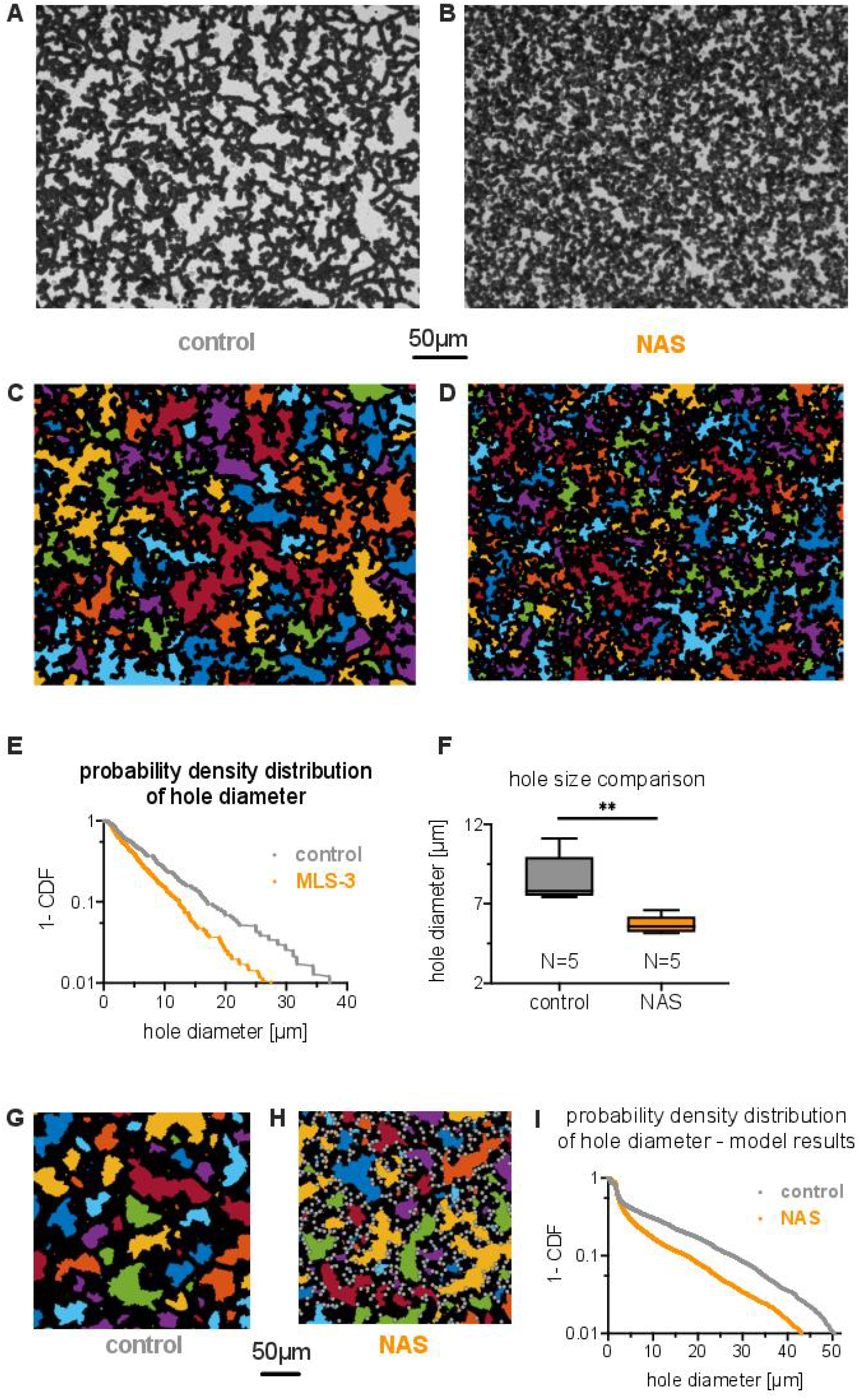
Characteristics of sedimenting erythrocyte aggregates in autologous plasma. A, B: Sedimented layer of erythrocytes in plasma for a control subject and a patient, on the bottom of a microscopy chamber after 2 h. Pictures are taken from the bottom of the well. A: healthy control donor and B: patient (MLS-3). The volume fraction (hematocrit) in the microscope slide well (inner diameter 5 mm, height 1.5 mm) was initially 0.33 % to reach a volume fraction close to 45 % at the bottom of the well after sedimentation. C, D: Same pictures as previously, but with the holes detected in the percolating network of erythrocytes highlighted with random colors. The 50 µm scale is valid for panels A to D. E: Example of distributions of the square root of the hole areas, or hole diameter, shown as complementary cumulative density functions. The linear curve reflects an exponential distribution with the mean hole diameter as a single scale parameter. This scale parameter was estimated by the maximum likelihood method. F: Statistical comparison of the scale parameters for control and NAS erythrocytes in autologous plasma. The scale parameters are normally distributed among each population, and a t-test between the two populations indicates a significant difference (*p* = 0.005). G: Simulation snapshot of healthy conditions with 100 % discoid erythrocytes shown in black color similar to experimental images. H: Simulated aggregate of a mixture of 80 % normal erythrocytes (black area) and 20 % acanthocytes (gray color) by area fraction. The scale bar is valid for panels G and H. I: Complementary cumulative density function of the square root of the hole areas from the simulations of healthy erythrocytes and erythrocyte-acanthocyte mixture exemplified in G and H.

**Figure 4.**
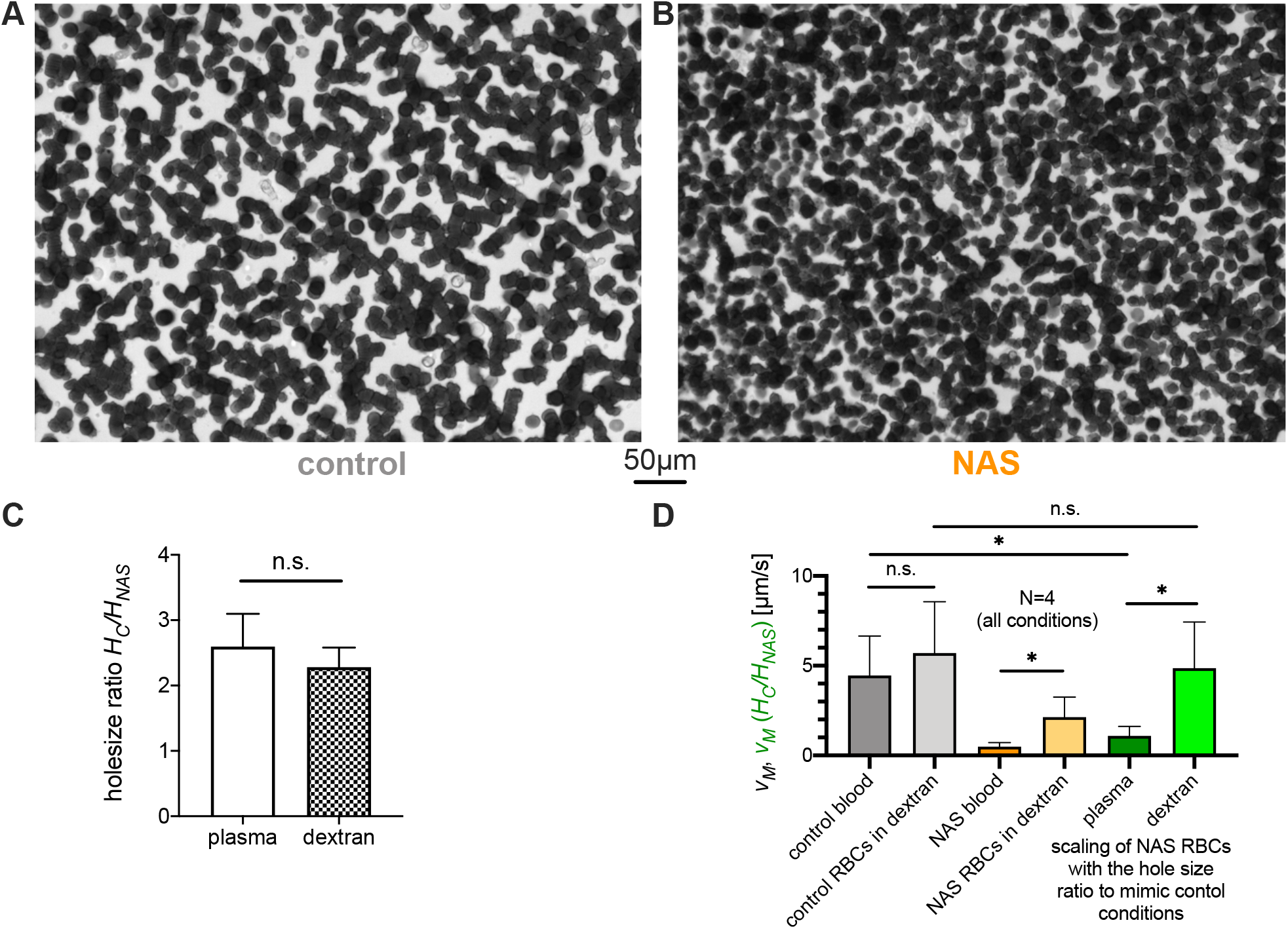
Characteristics of sedimenting erythrocyte aggregates in various suspension media. A,B: Sedimented layer of erythrocytes in 55 mg/ml dextran from A: a healthy control donor and B: an NAS patient. The characteristic area of the holes in the network is visibly smaller in the network of NAS patient’s erythrocytes. As the replacement medium, we used 70 kDa dextran diluted in PBS to a final mass concentration of 55 mg/ml. C: Measured ratio of mean hole size for the suspension media (autologous plasma and dextran diluted in PBS with a mass concentration of 55 mg/ml). The dextran concentration was empirically chosen in order to achieve a sedimentation speed as close as possible to the rate obtained in the plasma of the healthy samples. D: Characteristic (maximal) erythrocyte sedimentation velocities, with scaling based on aggregate geometries. The bars indicate the values of the characteristic erythrocyte sedimentation velocities. For each suspension medium, the patient scaling was obtained by multiplying the characteristic speed of each patient’s erythrocytes by the ratio of the characteristic hole areas, as justified by Eq. 3. While the scaled velocity in autologous plasma is significantly smaller than the characteristic speed of the control erythrocytes, the scaling lies within the same range when the autologous plasma is replaced by a dextran-based medium. n.s., not significant (p*>*0.05); * p*<*0.05.

#### Role of Erythrocytes in Sedimentation

The sedimentation of erythrocytes in blood plasma has been proposed, but was never established, to obey a so-called gel sedimentation regime^11,12^. Gel sedimentation is efficiently represented by several equations established for colloidal systems^13–16^. In particular, gel sedimentation can exhibit a delayed collapse^17,18^. Here, we apply the gel sedimentation model for sedimenting erythrocytes. This means that erythrocytes first experience only little sedimentation. Then, after a characteristic time on the order of a few minutes, some “cracks” or “channels” form within the aggregated structure of the erythrocytes, as suggested by a previous study^11^. We also observed this behavior in our samples (see Supplemental Fig. 5). It is worthwhile to notice that, due to volume conservation, the sedimentation of the erythocytes implies an upward flow of the plasma. The channel structures then enhance this upward flow of liquid. Here, we will apply for the first time some quantitative scaling based on these observations. Indeed, the maximal sedimentation speed *v*_*M*_ following volume conservation and Darcy’s law^15^ can be approximated as:

**Figure 5.**
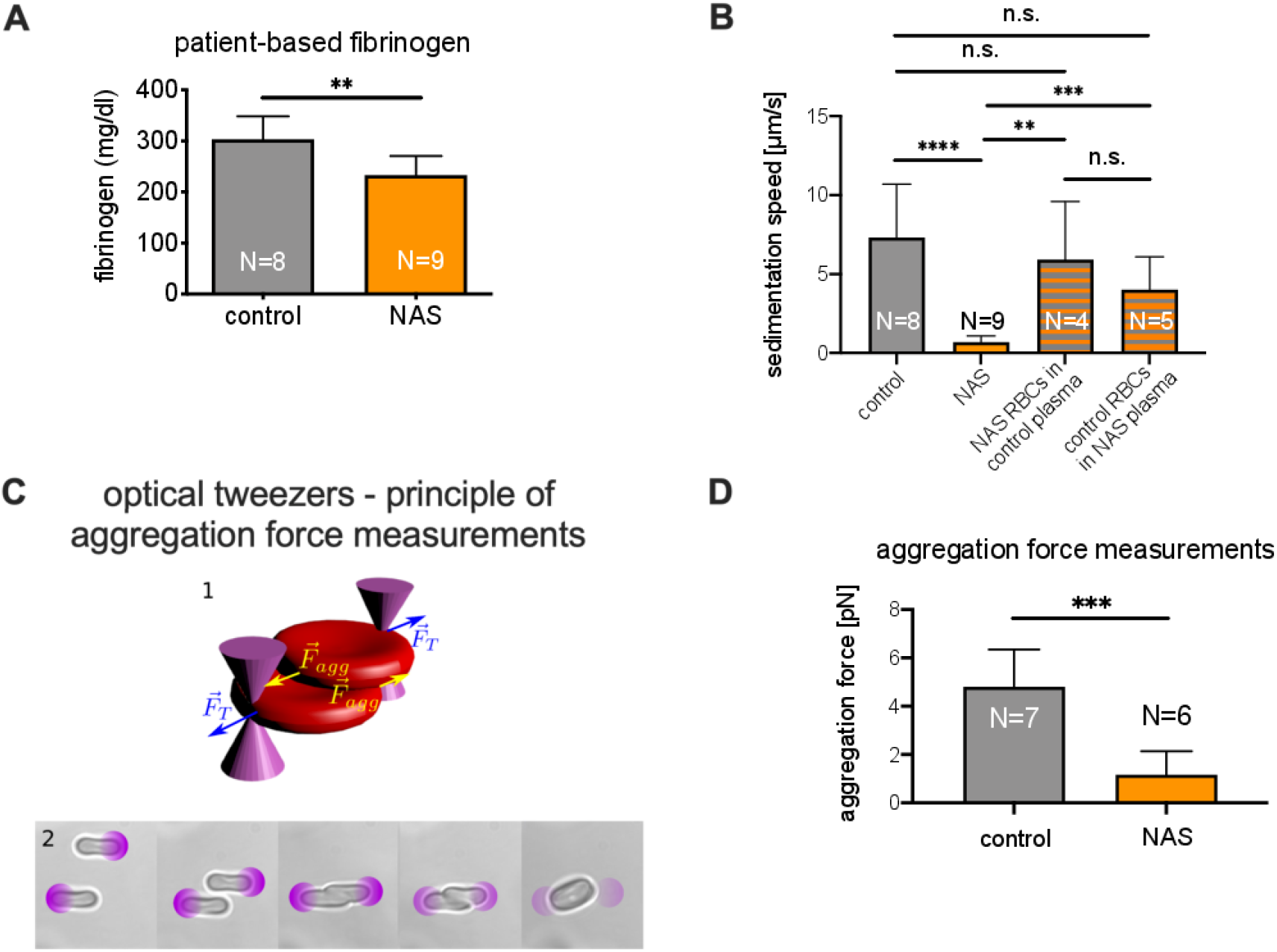
Measurements related to the role of plasma in sedimentation. A: Comparison of fibrinogen concentration. A significant difference consistent with the aggregation force difference is observed. B: Comparison of sedimentation velocities of control samples, NAS patient samples and samples with erythrocytes and plasma exchanged. C 1: Schematic of the forces acting on the erythrocytes trapped in optical tweezers. C 2: Microphotographs of the protocol for the measurement of the aggregating force. The purple circles show the location of external optical traps. From left to right: After being selected, erythorcytes were lifted 15 µm from the microscope slide by 4 optical traps, one for each erythrocyte extremity. The optical traps have known trapping forces. The erythrocytes are then brought into contact. At equilibrium, the two inner traps are removed. The optical force holding the cells is stepwise decreased, and the overlap distance tends to increase in the same manner. Finally, spontaneous aggregation overcomes the optical forces, and the erythrocytes escape the trap. The trapping force at which the cells aggregate is considered to be the aggregation force. D: Comparison of aggregation force. A significant difference between patients and controls was observed. n.s., not significant (p*>*0.05); ** p*<*0.01; *** p*<*0.001; and **** p*<*0.0001.

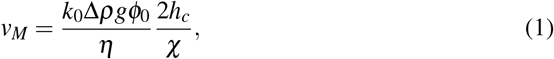

where *k*_0_ is the permeability of the aggregated erythrocyte structure, Δ*ρ* is the density difference between the erythrocytes and the suspending liquid, *ϕ*_0_ is the initial hematocrit, *η* is the liquid viscosity, *h*_*c*_ is the height of the channel, and *χ* is the characteristic radius of the zone where this channel influences the liquid flow^15^. Note that the permeability of the aggregated erythrocyte structure *k*_0_ is proportional to the characteristic area of the holes within the cross-section ⟨*H*⟩ of aggregated erythrocytes: *k*_0_ ∝ ⟨*H*⟩^13^. Therefore, the maximal sedimentation speed is also proportional to ⟨*H*⟩:

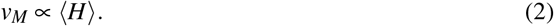

To estimate the hole size, we allowed the erythrocytes to settle in a microscope chamber such that the final hematocrit on the coverslip layer was close to the physiological value of 45 % (Fig. 3A,B). This, when observing the chamber from the bottom, allows a two-dimensional analysis of the hole size *H* based on microscopy images as outlined in Fig. 3C-E. The quantitative difference in average hole diameter (defined as the square root of the average hole area) is exemplified for one healthy control and one NAS patient by a probability density distribution (Fig. 3E). Statistical analysis of the hole diameters is given in Fig. 3F, revealing a larger hole size for the blood of healthy donors. One can also note that the total area covered by the erythrocytes is different between healthy controls and NAS patients (e.g., in Fig. 3A-D, the total area of the holes is 8.5 10^4^ µ*m*^2^ under control conditions but 6.2 10^4^ µ*m*^2^ for the patients). This is caused by the projection area of the cells despite the absolute hematocrit being equal between the groups. Under control conditions, almost all cells form rouleaux and hence contribute to a side projection, while acanthocytes and echinocytes have a larger projection area.

To better understand the effect of acanthocyte shape on the properties of sedimented aggregates (e.g., hole size), we performed a number of two-dimensional simulations with different compositions of healthy erythrocytes and acanthocytes; see example snapshots in Fig. 3G,H (and see Supplemental Data for simulation details). Figure 3I shows the corresponding cumulative density functions of hole diameter (defined as the square root of the hole area), which compare favorably to the experimental data in Fig. 3E. The simulations reveal that acanthocyte shape and rigidity effectively reduce aggregation interactions, as they cannot maximize locally aggregated membrane areas between cells, which is achieved by cell deformation in healthy erythrocytes. A lower effective aggregation between cells leads to smaller hole sizes, which we confirmed directly by reducing the aggregation strength in the suspension of healthy cells.

To further assess the role of erythrocytes in the reduced sedimentation rate and exclude the influence of individual plasma properties, sedimentation experiments with washed erythrocytes suspended in dextran (70 kDa) diluted in phosphate buffered saline (PBS) were performed. A dextran concentration of 55 mg/ml, which mimics erythrocyte aggregation in blood samples of healthy controls, was empirically selected (Fig. 4A,D). The aggregates formed in sedimented layers of erythrocytes in dextran solution (Fig. 4A,B) indeed looked very similar to the aggregates in autologous plasma (compare with Fig. 3A,B). The hole size ratio *H*_control_*/H*_NAS_ in the dextran solution was also very similar to that in plasma solution (Fig. 4C).

For further comparison, the maximum sedimentation speed *v*_*M*_ was measured for each ESR test. Under the assumption of a fixed hematocrit of 45 %, only the parameters *k*_0_ and Δ*ρ*, related to the properties of erythrocytes, can imply a difference in sedimentation speed. Standard blood parameters, i.e. hemoglobin concentration (Table 1, Supplemental Fig. 3A) and complete red blood cell count parameters, i.e. number of erythrocytes (Table 1, Supplemental Fig. 3B) and mean cell volume (MCV; Supplemental Fig. 3D) did not show any significant difference that could decrease the ESR of NAS patients through Δ*ρ*. However, MCHC (Supplemental Fig. 3C) was slightly increased in the NAS patients but, as previously mentioned, would then increase the ESR and therefore cannot explain the differences. Thus, different cases can be compared by scaling the observed maximum sedimentation speed with the hole size; see Eq. 2. If we denote patients with a subscript NAS and the healthy controls with C, then we obtain

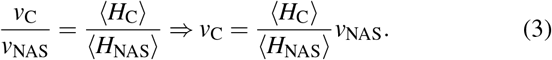

The *v*_*M*_ of control (*v*_*C*_) and NAS blood (*v*_*NAS*_) as well as the corresponding erythrocyte-dextran mixtures are shown in Fig. 4D (gray and orange columns, respectively). Additionally, the sedimentation speed was scaled according to Eq. 3. This scaling is plotted in Fig. 4D (green columns), along with the error bars corresponding to the standard deviation of the scaled speed measurements. The scaling makes the velocities comparable and indicates that the difference in the aggregate geometry as characterized by hole area accounts for the observed difference in sedimentation velocity when erythrocytes are suspended in a dextran-based medium. However, the scaling is not sufficient to explain the velocity ratio when the sedimentation is measured in autologous plasma, as shown in Fig. 1. This indicates that the aggregate geometry accounts for some fraction of the ASR slowdown, while the remaining fraction must correspond to other contributions, such as patient plasma composition.

#### Role of Plasma in the Sedimentation

As mentioned above, the total protein content likely contributes to the reduced ASR (Supplemental Fig. 4A). Therefore, we further analyzed plasma constituents, such as fibrinogen (Fig. 5A), albumin (Supplemental Fig. 4B), C-reactive protein (CRP, Supplemental Fig. 4C) and immunoglobulin G (IgG, Supplemental Fig. 4D). Since the last three did not show significant differences between NAS patients and healthy controls, we focused on fibrinogen, which was on average 23 % lower in the patients (*p* = 0.004).

To investigate the role of plasma in the increase in the ESR, we performed experiments in which the erythrocytes and plasma of patients were swapped with those of a healthy control with a matching blood group. We observed that healthy erythrocytes suspended in patient plasma sediment significantly faster than NAS erythrocytes in healthy plasma (which confirms the importance of erythrocyte properties). However, patient erythrocytes also sediment in healthy plasma significantly faster than in patient plasma, which is a clear sign that NAS plasma also slows sedimentation. Representative time traces are shown in Supplemental Fig. 6. A statistical presentation is given in Fig. 5B.

Previous reports have shown that an increase in fibrinogen concentration is related to an increased sedimentation speed and that fibrinogen has a larger impact on ESR than other proteins^19,20^. In particular, fibrinogen increases the interaction energy between erythrocytes^21^. To test whether this difference in interaction energy also occurs in NAS patients, we measured the aggregation forces between erythrocytes. To this end, we performed experiments using holographic optical tweezers, as shown in Fig. 5C. As depicted in Fig. 5D, the aggregation forces of erythrocytes from acanthocytosis patients in autologous plasma were significantly lower (*p* = 0.0005) than those from healthy controls.

The details of the physical mechanisms leading to delayed collapse due to the appearance of cracks in weakly aggregated colloids are still under investigation^11,12,16,22–25^. However, recent studies highlighted that the aggregation energy can modify the sedimentation speed in this regime, as well as the time of onset^22–25^. Therefore, the difference in plasma fibrinogen concentration is likely to explain the further difference in sedimentation speed between NAS patients and healthy controls.

## 3 Discussion and Conclusions

We herein not only present a decreased ESR in NAS but also conceptualize the biophysical reasons for this observation and highlight that measurement of the ESR needs to be implemented in routine clinical tests. Our measurements show that a decreased ESR can be used as an efficient biomarker to test for ChAc and MLS. For a sedimentation time of 2 h, we could (with our test tubes) determine a clear diagnostic threshold of 10 mm. In the context of a clinical workflow, the ESR (manually or automatedly determined) is a robust and cost-efficient parameter that could easily complement the error-prone acanthocyte count. Mutations can still be confirmed by molecular biology methods to differentiate among NASs (cp. Supplemental Data). Since the ESR also depends on the geometry of the tubes^26,27^, systems other than those used here may need their own standardization.

Conventionally, ESR is used to detect general inflammation, which is indicated by an increased ESR^28,29^; however, there is no conflicting overlap because the change is in the opposite direction in NAS. We would have liked to test whether the ASR correlates to the severity of the disease, but there is no validated index or score for NAS severity available. Similar to the acanthocyte count, ASR cannot discriminate between ChAc and MLS. Furthermore, we cannot make a statement about other NASs (such as pantothenate kinase-associated neurodegeneration (PKAN)), but we suppose a prolongation of the ESR also occurs in blood samples of patients with other diseases showing an increased number of acanthocytes.

The number of patients in our study may seem low (6 ChAc and 3 MLS patients), but given the ultrararity of these diseases—the estimated prevalence is less than 1 to 5 per 10^6^ inhabitants each^5^—the number of participants can well be regarded as acceptable. In particular, this holds true because with such a low number of patients, we were able to obtain significant results. Recent studies have identified promising drug targets, such as Lyn kinase, as a potential disease-modifying therapy for ChAc^5^. Considering the rarity and high variability of the natural history of the disease, there is an urgent need for a robust biomarker to achieve clinical trial readiness. The ASR might represent an ideal biomarker candidate in this context.

We have shown that both erythrocytes properties and blood plasma composition of NAS patients influence the ASR. Both components reduce the sedimentation speed, i.e. they are additive and provide a clear cut-off to differentiate from healthy individuals. This is to some extent surprising since the ESR is a measure that integrates numerous parameters, and its initial intended use as a parameter for reporting inflammation has lost importance to a certain degree within the last decades^30^ due to a lack of specificity.

Furthermore, we discussed physical mechanisms related to the erythrocyte and plasma properties that determine the ESR. Historical reports describe an altered aggregation behavior of acanthocytes^31^ and the influence of erythrocyte shape on the ESR^32^. Here, we applied the principles of colloidal physics to describe sedimenting erythrocytes in a quantitative manner. Thus, we clearly show that the geometric properties of erythrocyte aggregates influence the porosity of sedimenting aggregates, which has a direct impact on the sedimentation velocity. Furthermore, a modeling approach provides evidence that the shapes of the erythrocytes (acanthocytes and echinocytes) and their rigidity are the primary cause for the differences in aggregate geometry. Regarding the plasma composition, we highlighted a difference in fibrinogen concentration, which modifies the interaction energy between erythrocytes. While the detailed influence of this energy on the sedimentation process is still under active discussion, it is clearly a key feature in the sedimentation process.

Future investigations are required for the clinical exploration of the ASR in a broader context of acanthocyte-related diseases and its longitudinal value in individual patients and a deeper physical understanding of erythrocyte sedimentation. The clinical application comprises a broader test for neurodegenerative diseases as well as the verification of the ASR as a biomarker for NAS treatments as proposed above. For the mechanistic understanding of the process of erythrocyte sedimentation based on colloidal physics, we have provided an initial concept that also requires further research.

## 4 Materials and Methods

### Patients and Blood Sampling

Blood samples were collected from six ChAc and three MLS patients. Patients were being treated at the neurologic departments of Technische Universität Dresden and the University Hospital of the Ludwig-Maximilian-Universität Munich. Blood sample collection was approved by the ‘Ä rztekammer des Saarlandes’, ethics votum 51/18, and performed after informed consent was obtained according to the declaration of Helsinki. The diagnosis of ChAc and MLS was based on the clinical phenotype and was confirmed by detection of the VPS13A mutation and/or absence of the erythrocyte membrane chorein via Western blot (ChAc) or the detection of XK mutations (MLS)^33^.

Blood was taken in the morning during routine patient visits and immediately transported to Saarland University in Homburg and Saarbrücken for further analysis, which started 6 to 8 h after withdrawal. Because transportation can have a tremendous effect on blood parameters^34^, samples from relatives (if available) or the investigators were taken as transportation controls (in total, eight different individuals). This practice excluded patients and controls matched in terms of sex, age or weight. Furthermore, three patients came directly to Saarland University, which allowed the investigation of fresh samples. However, concerning the ASR, there was no significant effect between fresh and transported samples, as outlined in Supplemental Fig. 7.

### Measurements of the ESR

ESR/ASR measurements were performed in standard Westergren tubes (Dispette original, REF GS1500) with 200 mm height EDTA-blood. Color pictures of the tubes were automatically taken every minute for up to 50 h with a Canon EOS 500D camera to cover the whole range of sedimentation in every case. For each tube, a custom written MATLAB algorithm extracted the position of the interface at the top of the concentrated erythrocyte suspension with an accuracy of at least 0.1 mm. Since the displacement of the interface between two successive frames was close to the picture resolution, instantaneous sedimentation velocities were extracted from least squares fitting of the data points with cubic splines with 8 free interior knots, as allowed by the SLM (Shape Language Modeling) package^35^.

Further detailed measurement techniques are described in the Supplemental Data.

## Data Availability

Most of the data generated and/or analyzed during this study are present in the paper or the Supplementary Figures. Intermediate technical results or newly design analyses software is available upon reasonable request to the corresponding authors.

## Data Sharing Statement

The datasets generated and/or analyzed in the current study are available from the corresponding author on reasonable request.

## Acknowledgements

This work was supported by the Deutsche Forschungsgemeinschaft (DFG) in the framework of the research unit FOR 2688 “Instabilities, Bifurcations and Migration in Pulsatile Flows”, by the European Union Horizon 2020 research and innovation program under the Marie Skłodowska-Curie grant agreement No 860436 – EVIDENCE and by the Saarland University competitive funding (Forchungsausschuss). K.P. was supported by the Else Kröner Clinician Scientist Program (TU Dresden, Germany) and the Rostock Academy for Clinician Scientists (RACS, University of Rostock, Germany), A.H. is supported by the “Hermann und Lilly Schilling-Stiftung fü r medizinische Forschung im Stifterverband”. F.Y., T.J. and C.W. acknowledge funding from French German University (DFH / UFA). We are indebted to the Advocacy for Neuroacanthocytosis Patients (www.naadvocacy.org) for financial support for chorea Western blotting. We also gratefully acknowledge the computing time granted through JARA-HPC on the supercomputer JURECA at Forschungszentrum Jülich. We would like to thank Alexander Kihm and Dr. Hannes Glass for help in obtaining blood samples, their handling and transportation.We are grateful to Prof. Joan-Lluis Vives-Corrons and Jan de Zoeten for valuable discussion of the manuscript.

## Authors contribution

Ad. D. and K.P. suggested the joint study. K.P. and L.K. defined the project. K.P., A.H. and Ad.D. performed the diagnoses and identified the patients suitable for the study. Al.D., T.J., C.W. and L.K. designed the physical measurements. Al.D., T.J. and L.K. performed the ESR measurements and the microscope-based erythrocyte sedimenting experiments. Al.D. analyzed the sedimentation data and suggested the physical mechanisms. A.R. and G.S. performed the cell counts. F.Y. measured the aggregation forces. J.G. took responsibility for the measurement of the hematological parameters. D.A.F., A.K.D. and S.B. performed the numerical simulations. All authors interpreted the results and wrote or reviewed the manuscript.

## Supplemental Data

### Blood Smear Preparation and Evaluation

Blood smears were prepared as described by Storch et al. 2005^10^. Whole EDTA-blood was suspended in a 1:1 ratio in a 0.9 % sodium chloride solution containing 5 IU/ml heparin and incubated for 1 h at room temperature. Then, a drop of this solution was smeared onto a glass coverslip by using a second coverslip. Dried smears were stained according to the Pappenheim method. First, they were placed in May-Grünwald solution for 3-4 min and then washed with distilled water. This was followed by staining with Giemsa solution for 20 min followed by another washing step with distilled water. Images were visualized with an Olympus microscope (BX60, Olympus, Japan) with a 40 *×* Plan Fluor Objective and 2 *×* postmagnification and recorded with a CCD camera (DP73, Olympus, Japan). At least 400 cells were counted by visual inspection and classified into stomatocytes, discocytes, echinocytes, acanthocytes and ‘other’ cell shapes.

### Laboratory Parameters and Erythrocyte Preparation

When required, erythrocytes were washed by centrifugation (3000 rcf for 7 min). The supernatant was then removed and replaced by PBS before redispensing the erythrocytes. This process was repeated three times. During the last iteration, the PBS was replaced by an adequate preparation (e.g., dextran diluted in PBS). Complete erythrocyte count and plasma parameters were measured by standard methods in the Clinical Chemistry Laboratory of Saarland University Hospital (Homburg, Germany).

### Widefield Microscopy

Microscopic pictures were taken using an inverted microscope in transmission mode with a bright field and a blue filter to enhance erythrocyte contrast with the background. A 20 *×* objective was used to send pictures to a CCD camera. Samples were made by filling 30 µl well slides from Ibidi (µ-Slide 18 Well, Catalog no 81826, inner diameter 5 mm, interior height 1.5 mm) with a hematocrit of 0.33 %. The suspending liquid was either plasma from a control subject or a patient or a preparation of dextran in PBS. The volume fraction of 0.33 % was determined empirically to reach the percolation threshold, i.e., the smallest concentration that could produce percolating aggregates of erythrocytes on the lower side of the well when erythrocytes have sedimented. The final volume fraction at the bottom of the well after sedimentation is close to 45 %. Pictures of an area of 610 *×* 480 µm^2^ of the sedimented layer were recorded and then analyzed through a custom written MATLAB algorithm.

### Two-dimensional Simulations

Two-dimensional simulations were employed to study changes in the hole areas of aggregated mixtures of healthy erythrocytes and acanthocytes. Both healthy cells and acanthocytes were modeled as ring-like bead-spring chains, each consisting of 50 vertices, with a discocyte and star-like (having six rounded corners) shape, respectively. Even though the discocyte and star shapes have different areas, they have the same circumference, so that the aggregation interaction between cells did not depend on cell type. The total potential energy of the bead-spring chain model consists of three parts^36^. The first part is elastic energy, represented by nonlinear springs that connect neighboring vertices into a ring-like configuration. The second part accounts for the cellular bending resistance between two neighboring springs, with a bending modulus set to 50 *k*_*B*_*T*. Finally, the last contribution corresponds to an area constraint, which forces the discocyte shape of healthy erythrocytes with a fixed circumference. Note that acanthocytes are modeled as rigid bodies in simulations, as they have shown no significant deformation in previous experiments^37^.

The simulations were performed in a domain of size 300 *×* 300 µ*m*^2^ with periodic boundary conditions in both directions. Langevin dynamics with constant temperature were employed for time integration of the simulated system. Aggregation interactions between cells were modeled by the Lennard-Jones potential, *U* (*r*) = 4*ε*((*σ/r*)^12^ − (*σ/r*)^6^) for *r < r*_cut_ with *σ* = 0.3 µm and *r*_cut_ = 0.72 µm. The strength of the potential, *ε*, was selected from the range of 1.5 − 2.5 *k*_*B*_*T* for different simulations to reflect aggregation variability in experiments. The total hematocrit *ϕ* was set to 50 %. Two sets of simulations were performed, including one with only healthy erythrocytes and one with a mixture of 80 % healthy cells and 20 % acanthocytes (NAS case) by area fraction. Each set consisted of 11 simulations with different parameter values. Note that since the area of an acanthocyte is larger than that of a healthy cell, the total number of cells in the NAS case is smaller than that in the healthy case.

### Measurement of Aggregation Forces

The aggregation forces between erythrocytes were measured through holographic optical tweezers as previously described^38^. This is a versatile technique for measuring forces in the piconewton (pN) range and allows force measurement at a single-cell level. A single infrared beam Nd:YAG laser (1064 nm, 3 W, Ventus 1064, Laser Quantum, UK) is reflected by a parallel aligned nematic liquid crystal spatial light modulator (PAL-SLM, PPM X8267-15, Hamamatsu Photonics, Japan) and focused with a large numerical aperture oil immersion objective (60*×*, Nikon, Japan) in an inverted microscope (Nikon, TE 2000, Japan). The real-time manipulation of optical traps was made possible by imaging the sample with a CMOS camera (ORCA Flash 4.0 V3, Hamamatsu, Japan) and using a MATLAB routine to compute the phase. The optical force holding cells can be tweaked by varying the initial laser power. Measurements were carried out following the protocol depicted in Fig. 5C. For each measurement, two erythrocytes with a discocytic shape were manually selected. Each erythrocyte was held by two optical traps placed at their extremity. The lower erythrocyte was brought into contact with the upper erythrocyte by changing the vertical position of the traps holding the erythrocyte in steps of 1 µm/s. The overlapping contact length between the erythrocytes was initially set to 4.5 µm for all the measurements. The aggregation force corresponds to the force that is insufficient to counterbalance the spontaneous aggregation forces between the two erythrocytes. The same protocol was repeated to measure at least 7 pairs of cells in both the pathological and healthy samples. Optical tweezers were calibrated, and forces were determined based on Stokes’ law as in^39^.

## Statistical Analysis

Statistical analysis was performed in Prism8 (GraphPad Software, USA). All data sets were checked for normality of the distribution by the Shapiro-Wilk test. The ESR for controls, ChAc and MLS patients was evaluated by the Brown-Forsythe and Welch ANOVA tests. Further significance between two conditions was then performed by an unpaired t-test, except if otherwise stated in the figure legend. Significance p-values are abbreviated as n.s. (not significant) for p*>*0.05, * for p*<*0.05, ** for p*<*0.01, *** for p*<*0.001 and **** for p*<*0.0001. Correlation analysis was performed by simple linear regression, and the p-value provides the slope difference from zero.

### Confocal Microscopy and 3D Rendering

Approximately 5 µl of blood was placed into 1 ml of 0.1 % glutaraldehyde (Sigma-Aldrich, USA) solution in PBS to fix cells in the shape in which they occur in the circulation^40^. Erythrocytes (5 µl in 1 ml PBS) were stained with 5 µl of CellMaskTM Deep Red plasma membrane stain (0.5 mg/ml; Thermo Fisher Scientific, USA) for 24 h at room temperature. Then, the cells were washed 3 times by centrifugation at 4000 rcf for 5 min (Eppendorf Micro Centrifuge 5415 C, Brinkmann Instruments, USA) in 1 ml of PBS solution. After washing, the cells were resuspended in PBS and finally placed on a glass slide for confocal microscopy. Each labeled sample was placed between two glass slides for imaging (VWR rectangular coverglass, 24 *×* 60 mm^2^) by employing a piezo stepper for a 20 µm *z*-range. Confocal image generation was performed with a spinning disk-based confocal head (CSU-W1, Yokogawa Electric Corporation, Japan). Image sequences were acquired with a digital camera (Orca-Flash 4.0, Hamamatsu Photonics, Japan). A custom written MATLAB routine was used to crop single cells from each image and perform 3D reconstruction to enable visualization of the 3D shape of the cell. Each single-cell 3D image contained 68 individual planes with an extent of 100 by 100 pixels and a lateral (*x*/*y*) resolution of 0.11 µm/pixel. The piezo stepper had a minimal step width of 0.3 µm, defining the *z*-resolution. To compensate for the difference in resolution in the *x*/*y* and *z*-directions, we modified the *z*-scale by means of linear interpolation. Thus, the obtained *z*-stack had dimensions of 100 *×* 100 *×* 185 voxels. The image stacks were then passed to a custom written ImageJ script. By applying a fixed threshold for every image, the script binarized the confocal *z*-stack to retrieve the cell membrane as an isosurface.

---

**Supplemental Figure 1.**
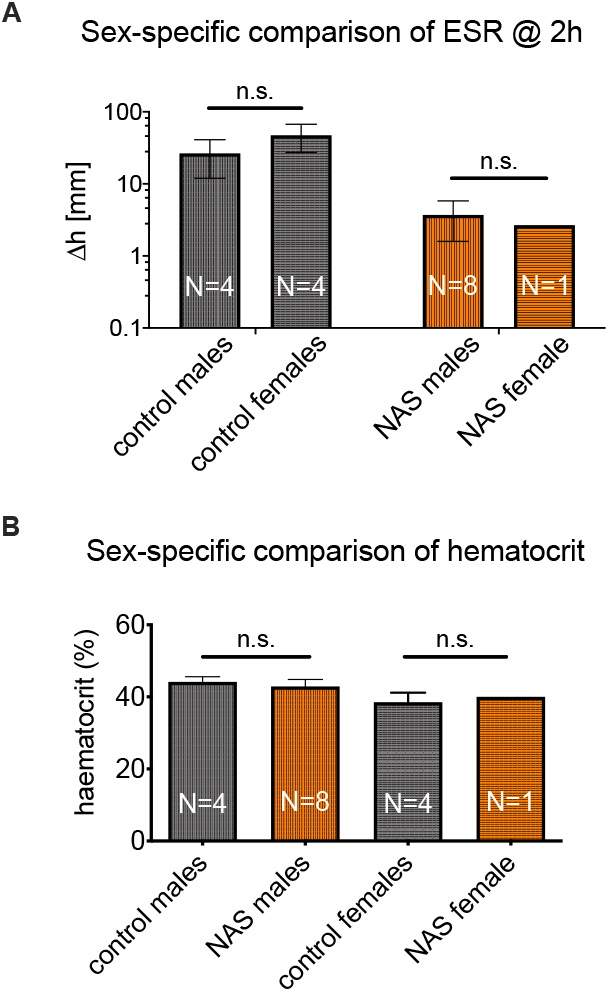
Sex-specific comparisons. A: Sex-specific comparison of the ESR after 2 h. In the control group, males and females are equally distributed, while in the patient group, the higher prevalence of the disease in males is reflected by the presence of only one female ChAc patient. We did not detect any significant sex-based difference between the controls and patients. B: Comparison of hematocrit. Since gender differences in the hematocrit are known, we compared this parameter separately for males and females between control and NAS blood samples. We did not find significant differences between healthy controls and NAS patients either in the gender-specific comparison as outlined here or when male and female subjects were pooled.

**Supplemental Figure 2.**
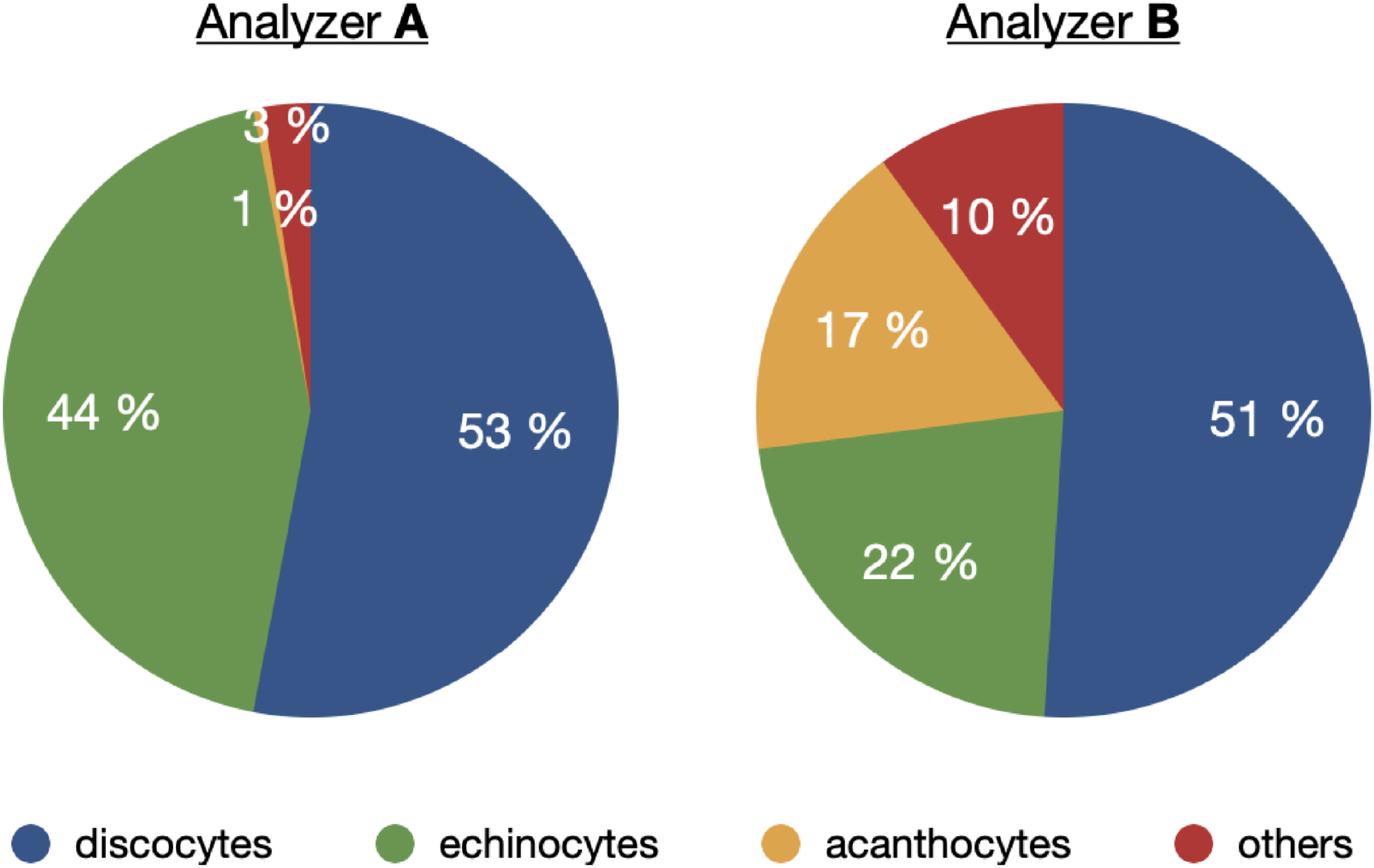
Analysis of blood smears. The blood smear from patient ChAc-5 was analyzed by two skilled staff members familiar with erythrocyte shapes. They were blinded to the intended comparison when they performed the counts. Approximately 1000 erythrocytes were counted by each analyzer. While the number of discocytes was consistent, the number of echinocytes, acanthocytes and other erythrocyte shapes (including stomatocytes and other cell shapes of known or unknown categories) varied tremendously. These differences reflect the uncertainty of acanthocyte counting based on blood smears.

**Supplemental Figure 3.**
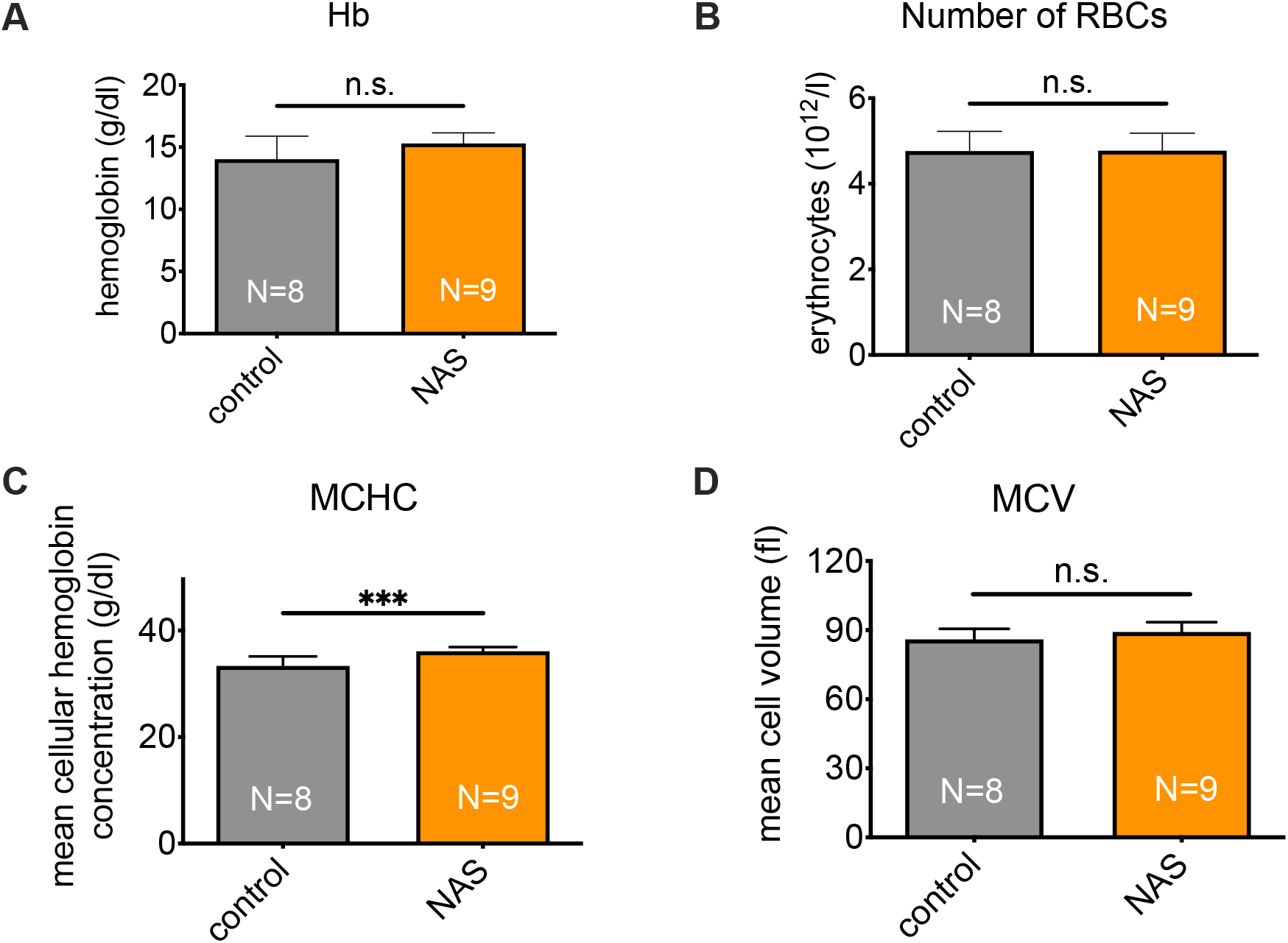
Comparison of red blood cells properties. A: Comparison of the hemoglobin concentration. For the hemoglobin (Hb) concentration, there was no significant difference between healthy controls and NAS patients. B: Comparison of the number of erythrocytes. For the number of erythrocytes (RBCs) per volume, there was no significant difference between healthy controls and NAS patients. C: Comparison of the mean cellular hemoglobin concentration. The mean cellular hemoglobin concentration (MCHC) was significantly increased in NAS patients. This is surprising since there were no differences in hematocrit (Supplemental Figure 1B), hemoglobin (panel A), number of erythrocytes (panel B) or mean cellular volume (panel D). The difference cannot be explained by the unequal gender distribution in the two groups. D: Comparison of the mean erythrocyte volume. For the mean cellular volume (MCV) of erythrocytes, there was no significant difference between healthy controls and NAS patients.

**Supplemental Figure 4.**
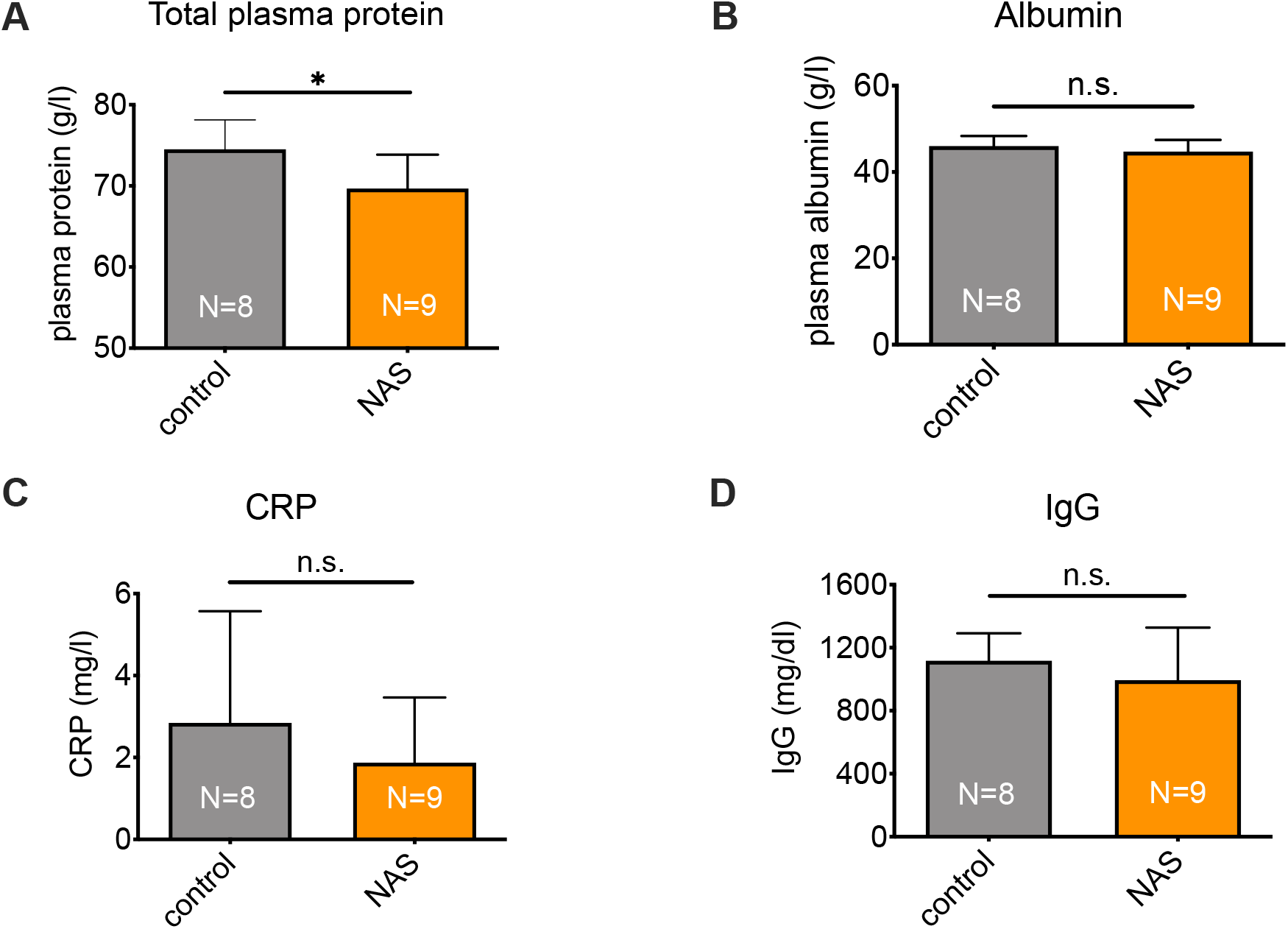
Comparison of plasma parameters. A: Comparison of total plasma protein. The total plasma protein was reduced in NAS patients (*p* = 0.023), providing an initial hint for the contribution of blood plasma to the reduced ESR. B: Comparison of albumin concentration. For the albumin concentration, there was no significant difference between healthy controls and NAS patients. C: Comparison of the C-reactive protein concentration. For the C-reactive protein (CRP) concentration, there was no significant difference between healthy controls and NAS patients. D: Comparison of the immunoglobulin G concentration. For the immunoglobulin G (IgG) concentration, there is no significant difference between healthy controls and NAS patients.

**Supplemental Figure 5.**
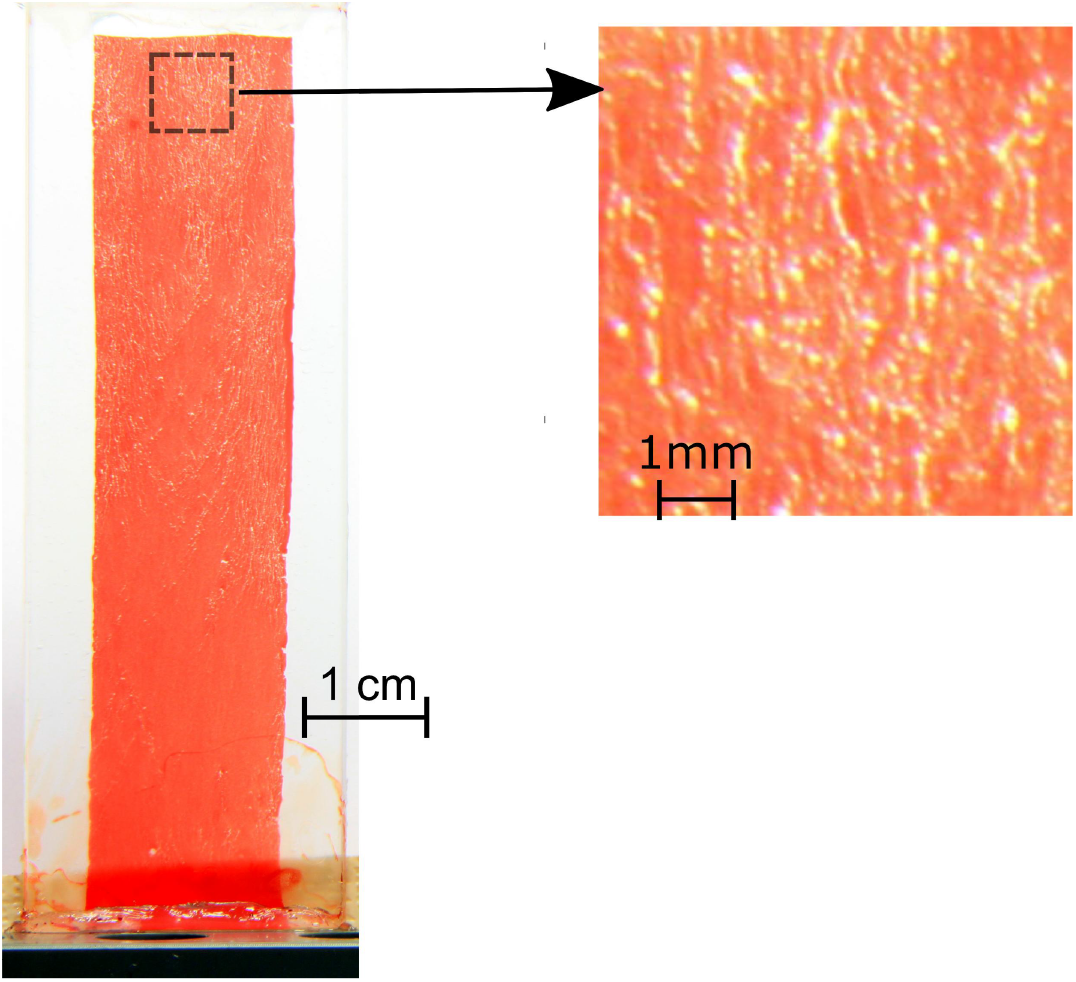
Channels that appear in sedimenting blood. The cell glass is approximately 100 µm thick, which makes the cracks visible to the naked eye. The square panel is a magnified view of the squared area in the rectangular picture. These channels are characteristic of the transient gel sedimentation regime of colloidal suspensions.

**Supplemental Figure 6.**
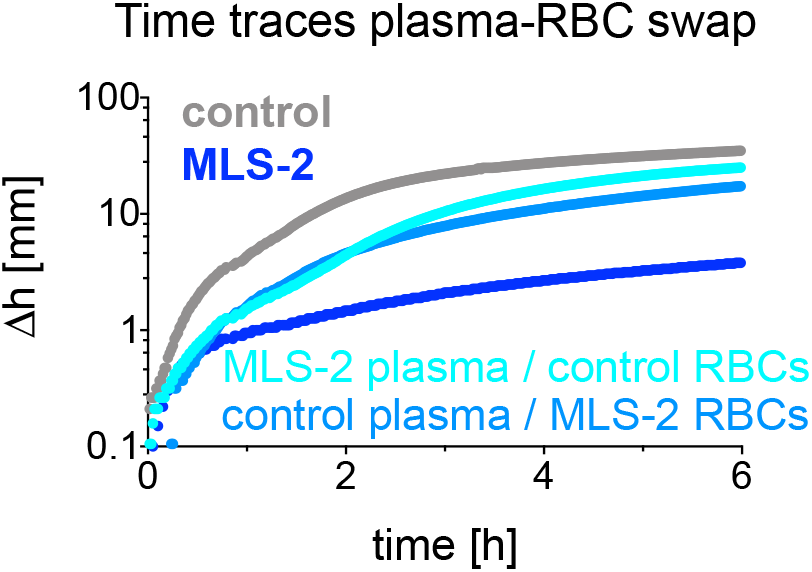
Example of time traces obtained by switching control and patient plasma. Erythrocytes from a control subject sediments slower in patient plasma. Erythrocytes from patient MLS-2 patient sediment faster in control plasma.

**Supplemental Figure 7.**
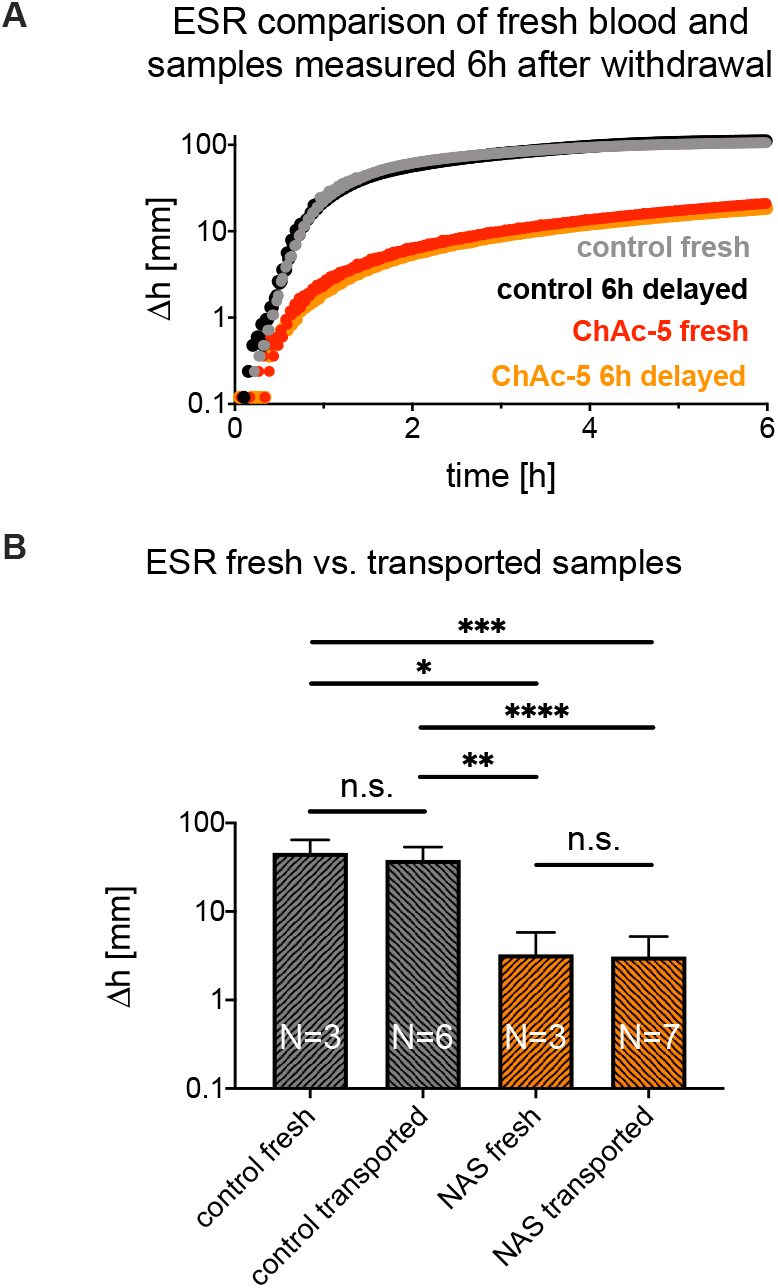
Comparison of fresh and delayed observations. A: Comparison of ESR time traces - fresh blood vs. 6 h after withdrawal. Blood samples from one healthy control and patient ChAc-5 were measured immediately after blood withdrawal and 6 h later. The delayed curves are almost indistinguishable from the fresh measurements.B: Comparison of the ESR after 2 h - fresh measurements vs. approximately 6 h of transportation. A subpopulation of the patients traveled to our laboratory in Saarbrücken to allow blood draws for immediate measurement. For the other patients, blood was collected at their local hospitals. We detected no difference in the ESR between fresh and transported blood for either the healthy controls or the NAS patients.

